# Disparities in SARS-CoV-2 exposure: evidence from a citywide seroprevalence study in Holyoke, Massachusetts, USA

**DOI:** 10.1101/2021.10.13.21264975

**Authors:** Wilfredo R. Matias, Isabel R. Fulcher, Sara M. Sauer, Cody P. Nolan, Yodeline Guillaume, Jack Zhu, Francisco J. Molano, Elizabeth Uceta, Shannon Collins, Damien M. Slater, Vanessa M. Sánchez, Serina Moheed, Jason B. Harris, Richelle C. Charles, Ryan M. Paxton, Sean F. Gonsalves, Molly F. Franke, Louise C. Ivers

## Abstract

**Background:** Seroprevalence studies are important tools to estimate the prevalence of prior or recent SARS-CoV-2 infections, identifying hotspots and high-risk groups and informing public health responses to the COVID-19 pandemic. We conducted a city-level seroprevalence study in Holyoke, Massachusetts, USA to estimate the seroprevalence of SARS-CoV-2 antibodies and risk factors for seropositivity.

**Methods:** We invited inhabitants of 2,000 randomly sampled addresses between November 5 and December 31, 2020. Participants completed questionnaires measuring sociodemographic and health characteristics, and COVID-19 exposure history, and provided dried blood spots for measurement of SARS-CoV-2 IgG and IgM antibodies. We calculate total and subgroup seroprevalence estimates based on presence of IgG antibodies using a Bayesian procedure that incorporates uncertainty in antibody test sensitivity and specificity. We account for clustering by household and weighting based on demographic characteristics to ensure estimates represented the city’s population.

**Findings:** We enrolled 280 households including 472 individuals. 328 underwent antibody testing. The citywide seroprevalence estimate of SARS-CoV-2 IgG was 13.1% (95%CI 6.9-22.3) compared to 9.8% based on publicly reported case counts. Seroprevalence was 16.1% (95%CI 6.2-31.8) among individuals identifying as Hispanic compared to 9.4% (95%CI 4.6-16.4) among those identifying as non-Hispanic white. Seroprevalence was higher among Spanish speaking households (21.9%; 95% CI 8.3-43.9) compared to English speaking households (10.2%; 95% CI 5.2-18.0) and among individuals living in high vulnerability areas (14.4%; 95% CI 7.1-25.5) compared to low vulnerability areas (8.2%; 95% CI 3.1-16.9).

**Interpretation:** The measured SARS-CoV-2 seroprevalence of IgG antibodies in Holyoke was only 13.1% during the second surge of SARS-CoV-2 in this region, far from accepted thresholds for “herd immunity.” Already vulnerable communities were at highest risk of prior infection. Implementation of local serosurveys in tandem with proactive public health interventions that address disparities in SARS-CoV-2 exposure are crucial to ensure at-risk communities have appropriate educational materials and access to vaccines, testing, and timely treatment.

**Funding:** The Sullivan Family Foundation, Harvard Data Science Institute Bias^2^ program, the US Centers for Disease Control and Prevention.

## Research in context

### Evidence before this study

The implementation of our seroepidemiologic study in Holyoke, Massachusetts December 2020 was informed by both the (1) variation in SARS-CoV-2 seroprevalence from surrounding communities and (2) lack of information on risk of SARS-CoV-2 infection by race and ethnicity status. Specifically, a serosurvey using convenience sampling of asymptomatic individuals in the predominantly Hispanic community of Chelsea, MA, during April 2020 demonstrated a high seroprevalence of 31.5%. This serosurvey was limited by non-representative convenience sampling that likely resulted in a biased estimate in addition to the use of a rapid lateral flow immunoassay with imperfect test accuracy. In July and August 2020, a university-related population and their household members in Massachusetts demonstrated a much lower seroprevalence between 4-5.3%. To our knowledge, these were the only two seroepidemiologic surveys in searchable databases at the time of this study. Globally, a recent systematic review and meta-analysis of SARS-CoV-2 serosurveys through December 2020 demonstrated significant regional variability in seroprevalence estimates and associated risk factors and highlighted similar methodological constraints as described above. Thus, our goal was to conduct a local serosurvey with the aim of informing community-tailored public health responses for at-risk groups, using population-representative sampling methods, and accounting for uncertainty in test performance characteristics.

### Added value of this study

We found that the seroprevalence of SARS-CoV-2 IgG antibodies was 13.1% during the second wave, far from acceptable levels of herd immunity, but also discrepant from the COVID-19 testing data from the city, which reported a prevalence of 9.6%. Hispanic and Latino/a individuals, individuals from Spanish-speaking households, and individuals living in areas with a high social vulnerability index had higher seroprevalence of SARS-CoV-2 IgG antibodies, indicating that already vulnerable communities were at a higher risk of prior infection. We utilized a novel Bayesian analytic approach to account for the sensitivity and specificity of the antibody test, while ensuring our study population reflected the population of Holyoke.

### Implications of all the available evidence

Our findings highlight the need for wide implementation of population-representative local serosurveys to ensure local details are not masked by findings at the state and national levels. In tandem with locally implemented serosurveys, proactive public health interventions will be crucial to respond to disparities in infection risk that will likely continue to propagate in the absence of further interventions.

## Background

Due to limitations in access to testing or because of the high frequency of asymptomatic infections, official reports have underestimated actual infection rates with severe acute respiratory syndrome coronavirus 2 (SARS-CoV-2), the virus that causes coronavirus disease 2019 (COVID-19), leaving public health officials with an incomplete picture of viral spread and associated risk factors.^1,2^ To address these limitations, seroepidemiologic prevalence studies (serosurveys) which measure the presence of anti-SARS-CoV-2 antibodies as a marker of prior infection have become an important public health tool to estimate incidence, identify risk factors and guide public health responses.^3^

A recent systematic review and meta-analysis of SARS-CoV-2 serosurveys through December 2020 demonstrated significant regional variability in seroprevalence estimates and associated risk factors.^3^ This reflects findings in the United States (US), where a national serosurvey across 10 sites during the first surge in 2020 documented a seroprevalence ranging between 1.0% in the San Francisco Bay area and 6.9% in New York City.^4^ In addition to regional variability, most SARS-CoV-2 serosurveys have suffered from several methodological constraints, predominantly related to non-representative sampling protocols and inaccuracies in serologic assays.

In the US, the Commonwealth of Massachusetts (MA), fueled by a super-spreader event of SARS-CoV-2, became one of the early epicenters of the pandemic.^5^ Variable local pandemic dynamics necessitate locally tailored responses, highlighting the importance of conducting local surveys to guide local public health responses to the SARS-CoV-2 pandemic.^3^

Holyoke is a post-industrial, majority Hispanic/Latino/Latina city in MA with high levels of socio-economic disadvantage. Based on 2018 census data, the poverty rate and median household income were 29.7% and $40,656, respectively, compared to 10.8% and $79,835 for MA.^6^ The city was disproportionately affected by the first surge of COVID-19 as evidenced by a high initial case count relative to the rest of the commonwealth.^7,8^ To fill the gap in our knowledge of city-level seroprevalence in the US and inform local public health responses, we conducted a serosurvey of the city of Holyoke, MA, during the second surge of SARS-CoV-2 cases between November and January 2021. Measurement of antibodies against SARS-CoV-2 was paired with questionnaires to identify household and individual-level risk factors associated with prior infection with SARS-CoV-2.

## Methods

### Study Design and Sampling

We conducted a representative, cross-sectional household-based seroepidemiologic SARS-CoV-2 survey in the City of Holyoke, MA. We obtained an official list of the 17,828 addresses in the city’s 11 census tracts from the city records. We identified and removed listings corresponding to congregate living facilities, vacant buildings, duplicate entries, and empty listings. The remaining addresses were considered eligible for sampling. We then randomly sampled 2,000 addresses from this final list.

### Participant Recruitment

Participants were enrolled from November 5, 2020, through December 31, 2020. The study team mailed an invitation letter to sampled addresses that contained a QR code and a unique ID for an online survey hosted on REDCap (Research Electronic Data Capture) at Massachusetts General Hospital. Among selected addresses, all individual household members aged ≥ 6 months and residing at the address were eligible for enrollment. A household was defined as a group of persons who slept under the same roof most nights. Households were given a period of approximately five days to respond by either taking the online survey or contacting the study team. Households could opt-out of future communication by calling a phone number. Following that period, data collectors made reminder follow-up calls, where participants had the option of completing the survey over the phone, and conducted home visits to follow-up with households that lacked a telephone number, were unresponsive to calls, or requested a visit from the team. To raise awareness about the study and increase community engagement, the study team distributed fliers and made announcements via Facebook and local media in both English and Spanish.

Following the first month of recruitment, we mailed a second wave of invitation letters to the originally sampled households, excluding ineligible addresses and households that had either already completed their surveys or opted out of the study. Data collectors made follow-up phone calls to households with incomplete surveys and those who had not mailed back their samples.

### Data Collection

The survey tool consisted of an eligibility and informed consent form, one household-level survey, and individual adult and child surveys for each consenting adult and child household member. Surveys included questions regarding sociodemographic characteristics, occupation and employment history, clinical history, symptoms, and COVID-19 testing, and exposure history. Upon completion of these surveys, blood test kits were either mailed to participant addresses or physically provided during home visits. We provided a $25 gift card to each household in which at least one person completed the entire study.

### Sample Collection, Transportation and Laboratory Testing

Test kits contained supplies and instructions (in English and Spanish) for self-collection of dried blood spot (DBS) samples. Individuals were instructed to perform a pinprick on the finger using a lancet and applying it to Whatman^®^ Protein Saver 903 filter paper collection cards (https://www.cytivalifesciences.com). Once obtained, samples were return mailed using a preaddressed envelope to Massachusetts General Hospital (Boston, MA, USA) where they were stored at −20°C with desiccant until tested. DBS sampling has been validated for use in antibody testing of SARS-CoV-2 and other pathogens.^9,10^

DBS were eluted and tested for the presence of SARS-CoV-2 IgG and IgM receptor-binding domain of the spike protein of SARS-CoV-2 using a quantitative ELISA previously developed and validated using reverse transcriptase polymerase chain reaction (RT-PCR)-positive mild and severe SARS-CoV-2 infections and pre-pandemic samples at Massachusetts General Hospital.^11–13^ The test specificity and sensitivity estimates were respectively 99.5% (95% CI 99.0-99.8) and 70.6% (95% CI 61.2-79.3%) for IgG antibodies and were calculated using samples from the Boston area. Further details provided in Section 2 of the Supplementary Materials.

### Statistical Analysis

Our main SARS-CoV-2 seroprevalence estimate was the proportion of the population that had IgG antibodies detected as this aligns with prior studies. We also calculated seroprevalence estimates with corresponding 95% credible intervals for the following combinations of IgG and IgM antibody positivity: IgG or IgM, IgG only, IgM, and IgM only.

We used a modified version of the Bayesian procedure proposed by Stringhini et al.^14^ to calculate prevalence estimates with corresponding 95% credible intervals. The procedure accounted for uncertainty in the antibody test sensitivity and specificity. Within the procedure, a random intercept was used to account for clustering by household and weighting was applied to ensure the seroprevalence estimates were representative of the population of the City of Holyoke. Details on the procedure are provided in Section 1 of the Supplementary Materials.

A raking procedure was used to construct weights based on the distribution of age, race and ethnicity, gender, and census tract in the City of Holyoke from the 2019 American Community Survey (ACS) (Table 1). Notably, gender is reported as the percent of “Female persons” in Holyoke in the ACS survey results, such that we only have two categories for gender: Female and non-Female, which includes male, transgender, and non-binary persons. For the purposes of constructing weights, we collapsed sparse categories of race and ethnicity into a “Grouped category”. The census tracts were also collapsed into “high vulnerability” and “low vulnerability” groups based on the social vulnerability index (SVI).^15^ High vulnerability was defined as having an SVI greater than the 75^th^ percentile of census tracts in Massachusetts – 9 of the 11 census tracts were considered high.

**Table 1.** Unweighted demographic characteristics of survey participants compared with 2019 American Community Survey estimates for the city.

| Characteristic |  | Responded to Survey |  | Provided Antibody Sample |  | Holyoke City<sup>1</sup> |  |
| --- | --- | --- | --- | --- | --- | --- | --- |
|  |  | N = 472 | % | N = 328 | % | N = 40241 | % |
| **Age group (years)** | **Median (IQR)\*** |  |  |  |  |  |  |
| 0 – 19 | 14 (8,16) | 50 | 10.6 | 27 | 8.2 | 10406 | 25.8 |
| 20 – 44 | 33 (27, 37.25) | 130 | 27.5 | 76 | 23.2 | 14335 | 35.7 |
| 45 – 59 | 53 (50.25, 56) | 135 | 28.6 | 94 | 28.7 | 7607 | 18.9 |
| 60 – 84 | 68 (64, 73) | 149 | 31.6 | 123 | 37.5 | 7019 | 17.4 |
| 85 and over | 88 (86.5, 89.25) | 8 | 1.7 | 8 | 2.4 | 874 | 2.2 |
| **Gender** |  |  |  |  |  |  |  |
| Female |  | 263 | 55.7 | 180 | 54.9 | 20747 | 51.6 |
| Male |  | 200 | 42.4 | 139 | 42.4 | 19494 | 48.4 |
| Transgender woman |  | 1 | 0.2 | 1 | 0.3 | -<sup>+</sup> | - |
| Transgender man |  | 0 | 0.0 | 0 | 0 | - | - |
| Non-binary |  | 7 | 1.5 | 7 | 2.1 | - | - |
| Prefer not to answer |  | 1 | 0.2 | 1 | 0.3 | - | - |
| Other |  | 0 | 0.0 | 0 | 0 | - | - |
| **Race/Ethnicity** |  |  |  |  |  |  |  |
| Hispanic or Latino/Latina |  | 126 | 26.7 | 66 | 20.1 | 21704 | 53.9 |
| Non-Hispanic or Non-Latino/Latina |  | 346 | 73.3 | 262 | 79.9 | 18537 | 46.1 |
| White |  | 311 | 65.9 | 239 | 72.9 | 16636 | 41.3 |
| Black or African American |  | 10 | 2.1 | 5 | 1.5 | 1162 | 2.9 |
| Asian |  | 7 | 1.5 | 5 | 1.5 | 239 | 0.6 |
| AIAN<sup>2</sup> |  | 1 | 0.2 | 1 | 0.3 | 78 | 0.2 |
| NHPI<sup>3</sup> |  | 0 | 0.0 | 0 | 0.0 | 0 | 0.0 |
| Other Race |  | 6 | 1.3 | 5 | 1.5 | 3 | 0.0 |
| Two or more races |  | 6 | 1.3 | 2 | 0.6 | 419 | 1.0 |
| Prefer not to answer |  | 5 | 1.1 | 5 | 1.5 | - | - |

<sup>1</sup>U.S. Census Bureau (2019). American Community Survey, Demographic and Housing Estimates. Table DP05.<sup>2</sup>AIAN = American Indian or Alaskan Native.<sup>3</sup>NHPI = Native Hawaiian and Other Pacific Islander.
\*Median (IQR) for the population of 328 individuals providing an antibody sample.
<sup>+</sup> - indicates that this data was not available in the American Community Survey.

We compare seroprevalence estimates and 95% credible intervals (CI) across subgroups. To investigate disparities in known risk factors for SARS-CoV-2 infection, we report sociodemographic and clinical factors stratified by race/ethnicity. Analyses were conducted in R V4.0.0 using *survey* and *rstan* packages.^16^

#### Ethics

Informed consent was obtained from adults 18 years or older. Informed parental consent and assent was obtained for children ages 14 – 17. Parental consent was obtained for children under the age of 14, with documented verbal assent by the caregiver sought for minors between the ages of 7 – 13. This protocol was reviewed by the Mass General Brigham Human Research Committee Institutional Review Board (Protocol ID: 2020P002560, November 2^nd^, 2020).

### Results

#### Study population

From the final list of 17,204 addresses, we randomly sampled 2,000 and mailed invitation letters and followed up recruitment as described above. 280 households (14%) with 472 individuals agreed to participate and completed household and individual-level questionnaires. Figure 1 demonstrates a complete flow diagram of participant progress through study phases. Mean household size was 2.28 individuals (SD: 1.2). Within participating households, an average mean of 1.77 household members (SD 1.03) consented to participate. Of these, 197 households (70.4%), 330 individuals (69.9%) completed and returned a DBS sample for analysis. A total of 328 samples from 195 households were analyzed. The mean household size for households with individuals that provided a DBS sample and were analyzed was 2.09 individuals (SD: 1.13), and the mean number of individuals per household that consented to participate was 1.71 (SD 0.96).

**Figure 1.**
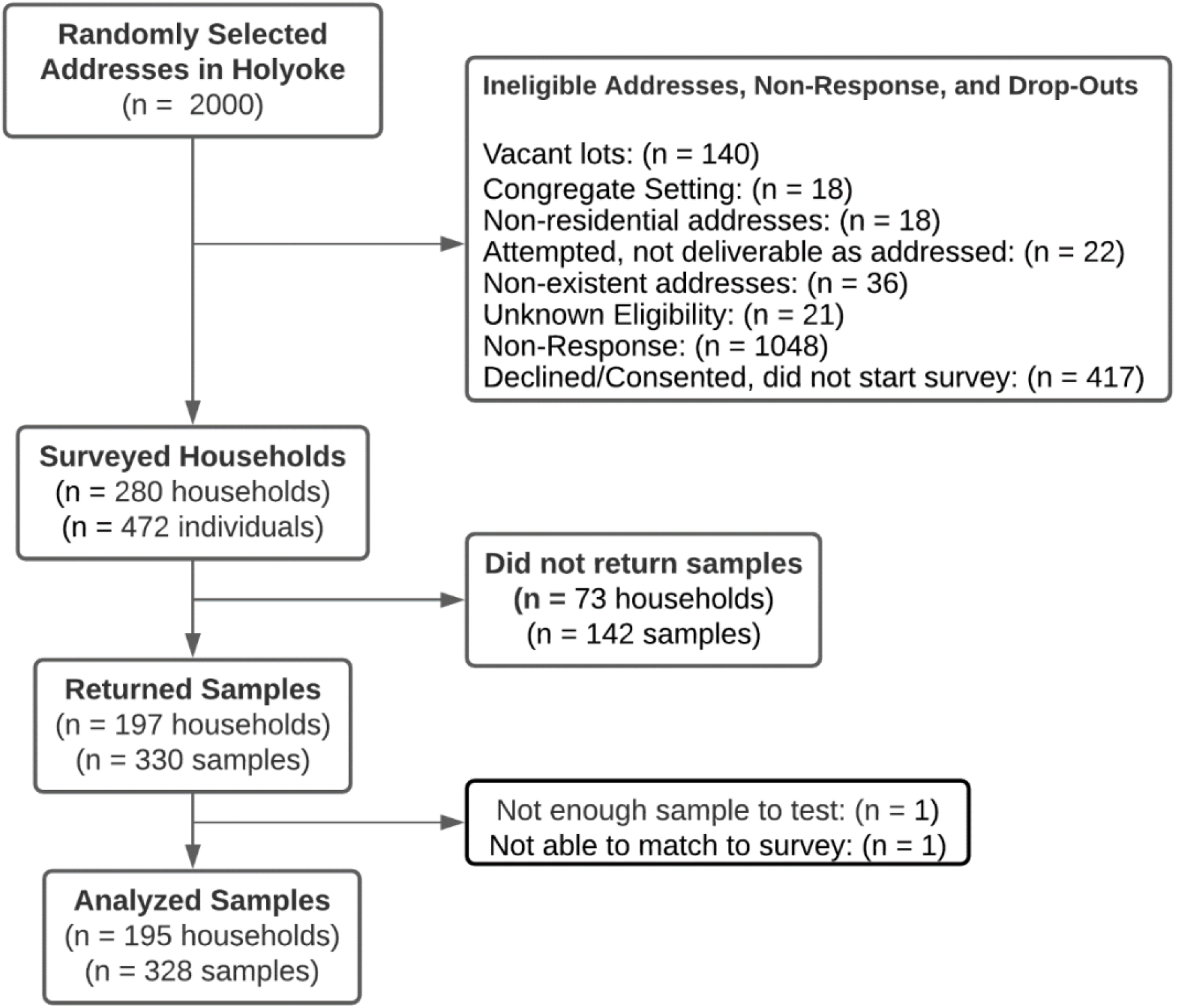
Study Participant Flow Diagram.

Among the 280 households that completed the survey, 37 reported hospitalization of a household member since February 2020 (13.3%, 1 missing response). Only one of these hospitalizations was reported to be a result of COVID-19. There were 2 reported deaths amongst the households since February 2020 among 277 households (3 missing responses). 25 of 471 respondents (5.31%, 1 missing response) described being diagnosed by a healthcare provider with pneumonia or other respiratory infection (may include COVID-19 diagnosis) since February 2020. 20 of the 468 respondents (4.3%, 4 missing response) reported testing positive for COVID-19 at least once.

Demographic characteristics including gender, age group and race/ethnicity breakdown for the entire study population is listed in Table 1. Individuals from younger age groups and individuals identifying as Hispanic or Latino/Latina were underrepresented in the study population relative to the population of Holyoke.

#### Citywide Seroprevalence of SARS-CoV-2 Antibodies

Of 328 individual samples tested, 27 individuals from 20 households were positive for SARS-CoV-2 IgG or IgM antibodies; after adjusting for clustering, differential response rates and imperfect test sensitivity, this corresponded to a citywide seroprevalence estimate of 13.6% (95%CI: 6.7-23.7). 25 individuals were positive for SARS-CoV-2 IgG antibodies, corresponding to an adjusted citywide seroprevalence estimate of 13.1% (6.9-22.3%). Seroprevalence estimates for individuals with IgG only, IgM only, and IgG and IgM are listed in Table 2. In the Supplemental Materials, we provide seroprevalence estimates under various sensitivity estimates ranging from 60.2% to 78.8%, corresponding to overall IgG seroprevalence estimates ranging from 15.3% to 11.7%, respectively.

**Table 2.** Seroprevalence by antibody positivity profile.

| **Characteristic** | **No.**<br>**tested** | **No.**<br>**Positive** | **Seroprevalence**<br>**% (95% CI)** |
| --- | --- | --- | --- |
| IgG | 328 | 25 | 13.1 (6.9-22.3) |
| IgG only | 328 | 18 | 7.8 (3.4-16.4) |
| IgM | 328 | 9 | 11.0 (2.3-25.9) |
| IgM only | 328 | 2 | 0 (0-5.0) |
| IgG or IgM | 328 | 27 | 13.6 (6.7-23.7) |

#### Seroprevalence of SARS-CoV-2 Antibodies by Sociodemographic Characteristics

Seroprevalence estimates were calculated for the following subgroups: age groups, gender, race/ethnicity, social vulnerability index, primary language spoken in the household, and other sociodemographic characteristics listed in Table 3. Seropositivity varied across multiple subgroups, though confidence intervals tended to be wide. The seroprevalence estimate was highest among individuals 20-44 years old (17.6%; 95%CI 7.5-32.4) and decreased with age. Likewise, the seroprevalence estimate was higher at 16.1% (95%CI: 6.2-31.8) among individuals identifying as Hispanic or Latino/Latina compared to a seroprevalence estimate of 9.4% (95%CI: 4.6-16.4) among individuals identifying as white. The seroprevalence estimate among Spanish speaking households was 21.9% (95% CI 8.3-43.9) compared to 10.2% (95% CI 5.2-18.0) among English speaking households. Individuals living in high vulnerability areas (14.4%; 95% CI 7.1-25.5) had a higher seroprevalence estimate than individuals living in low vulnerability areas (8.2%; 95% CI 3.1-16.9).

#### Seroprevalence of SARS-CoV-2 Antibodies by Medical, Symptom and Exposure History

We investigated seroprevalence by medical conditions and COVID-19 symptoms, testing, and exposure history (Figure 2). Notably, the seroprevalence estimate of individuals reporting any of the listed symptoms since February 2020 was 18.6% (95%CI 9.6-31.4) compared to a seroprevalence of 5.6% (95%CI: 1.0-15.3) among individuals reporting no symptoms since that time. There was also variation in seroprevalence estimate by the type of exposure to individuals with confirmed COVID-19. The seroprevalence among individuals reporting an exposure to a household member was 72.4% (95%CI: 32.6-99.7), while only 21.3% (95%CI: 8.0-41.5%) among those reporting an exposure to a non-household member and 5.8% (95%CI: 1.9-13.4%) among those reporting no known exposure to someone with COVID-19.

**Table 3.** Seroprevalence^1^ by sociodemographic characteristics.

| Characteristic (N if not 328) | N | n | Weighted<sup>4</sup> |
| --- | --- | --- | --- |
|  |  |  | Seroprevalence,<br>% (95% CI) |
| **Age groups (years)** |  |  |  |
| 0-19 | 27 | 2 | 11.2 (1.9-32.9) |
| 20-44 | 76 | 10 | 17.6 (7.5-32.4) |
| 45-59 | 94 | 6 | 9.8 (3.4-21.1) |
| 60 and above | 131 | 7 | 8.9 (3.6-17.4) |
| **Gender** |  |  |  |
| Female | 180 | 15 | 14.4 (6.7-26.0) |
| Grouped categories (Male or Non-binary) | 148 | 10 | 11.6 (5.0-22.4) |
| **Race/Ethnicity** |  |  |  |
| non-Hispanic, White | 239 | 16 | 9.4 (4.6-16.4) |
| Hispanic or Latino/Latina | 66 | 8 | 16.1 (6.2-31.8) |
| non-Hispanic, Grouped categories | 23 | 1 | 7.3 (0.7-24.4) |
| **Social Vulnerability Index** |  |  |  |
| High | 185 | 16 | 14.4 (7.1-25.5) |
| Low | 143 | 9 | 8.2 (3.1-16.9) |
| **Primary language spoken in the household** |  |  |  |
| English | 275 | 18 | 10.2 (5.2-18.0) |
| Spanish | 36 | 7 | 21.9 (8.3-43.9) |
| Multi-lingual/Other | 16 | 0 | 3.9 (0.1-27.0) |
| Missing | 1 | 0 |  |
| **Highest education level (N = 305)\*** |  |  |  |
| Some high school or less | 22 | 2 | 12.5 (2.2-34.8) |
| High school/GED or some college | 95 | 8 | 15.2 (5.9-29.0) |
| Associate or bachelor's degree | 101 | 7 | 12.6 (4.8-24.7) |
| Master's doctorate or professional degree | 80 | 6 | 11.7 (4.0-23.6) |
| Missing | 7 | 0 |  |
| **Employment status on February 1st, 2020 (N = 305)\*** |  |  |  |
| Working | 181 | 19 | 18.1 (9.7-29.6) |
| Not Working | 94 | 2 | 3.9 (0.5-12.7) |
| Other | 29 | 2 | 11.5 (1.1-32.0) |
| Missing | 1 | 0 |  |
| **Worked outside home during "stay at home" order (N=305)\*** |  |  |  |
| No | 211 | 13 | 10.8 (4.8-20.1) |
| Yes | 93 | 10 | 19.5 (9.4-33.8) |
| Missing | 1 | 0 |  |
| **Worked as a health worker in a healthcare setting (N=93)\*+** |  |  |  |
| No | 75 | 7 | 13.1 (4.4-27.7) |
| Yes | 18 | 3 | 20.7 (4.8-53.4) |
| **Workplace offered paid sick leave, work-from-home, overtime, or hazard pay during Mar – June (N=93)<sup>1\*</sup>** |  |  |  |
| None of the above | 31 | 4 | 15.3 (3.2-38.7) |
| 1 – 2 of the above | 56 | 5 | 13.6 (4.5-30.9) |
| 3 – 4 of the above | 6 | 1 | 8.4 (0.3-45.1) |
| **Travel since February 1st, 2020 (N = 328)** |  |  |  |
| No | 114 | 8 | 12.5 (4.5-25.2) |
| Yes, to a different city | 117 | 5 | 6.9 (1.4-16.2) |
| Yes, to a different state | 88 | 8 | 10.8 (3.4-26.5) |
| Yes, to a different country | 7 | 4 | 55.5 (14.2-98.0) |
| Missing | 2 | 0 |  |
| **Annual household income (N = 328)** |  |  |  |
| Less than \$15,000 | 21 | 6 | 33.8 (10.6-66.9) |
| \$15,000 - \$50,000 | 69 | 1 | 2.0 (0-14.7) |
| \$50,000 - \$80,000 | 64 | 3 | 7.4 (0.3-22.4) |
| \$80,000 - \$160,000 | 94 | 9 | 15.8 (6.3-29.2) |
| >\$160,000 | 24 | 4 | 13.9 (2.7-38.3) |
| Prefer not to answer | 53 | 2 | 8.2 (0.1-26.0) |
| Missing | 3 | 0 |  |
| **Home type (N = 328)** |  |  |  |
| Single Family | 189 | 11 | 8.4 (3.6-16.1) |
| Multi-family | 67 | 5 | 14.0 (4.2-29.4) |
| Apartment or condominium | 67 | 8 | 15.9 (5.8-33.4) |
| Other | 3 | 1 | 23.1 (2.2-69.8) |
| Missing | 2 | 0 |  |
| **Rent or own the home (N = 328)** |  |  |  |
| Someone in the household owns the home | 235 | 16 | 12.1 (5.6-21.5) |
| We have a landlord | 88 | 8 | 13.8 (5.0-29.4) |
| Missing | 5 | 1 |  |
| **Number of people living in the household** |  |  |  |
| One | 65 | 2 | 4.7 (0.3-14.7) |
| Two | 130 | 11 | 13.9 (6.1-26.0) |
| Three | 73 | 3 | 3.0 (0.0-17.1) |
| Four or more | 60 | 9 | 25.8 (9.2-50.6) |

<sup>1</sup>Seroprevalence for demographics groups based on IgG antibody positivity (i.e., at least IgG positive).<sup>2</sup>N refers to the total numbers of individuals in each category.<sup>3</sup>n refers to the total number of individuals seropositive for IgG in each group.<sup>4</sup>Weights were computed as the inverse probability of selection and adjusted so that the marginal distribution of age group, gender, race/ethnicity, and social vulnerability index of the sample agreed with population estimates.
\*Among adult respondents only.
\*Among individuals that responded “Yes” to working outside the home during the “stay at home” order.

**Figure 2.**
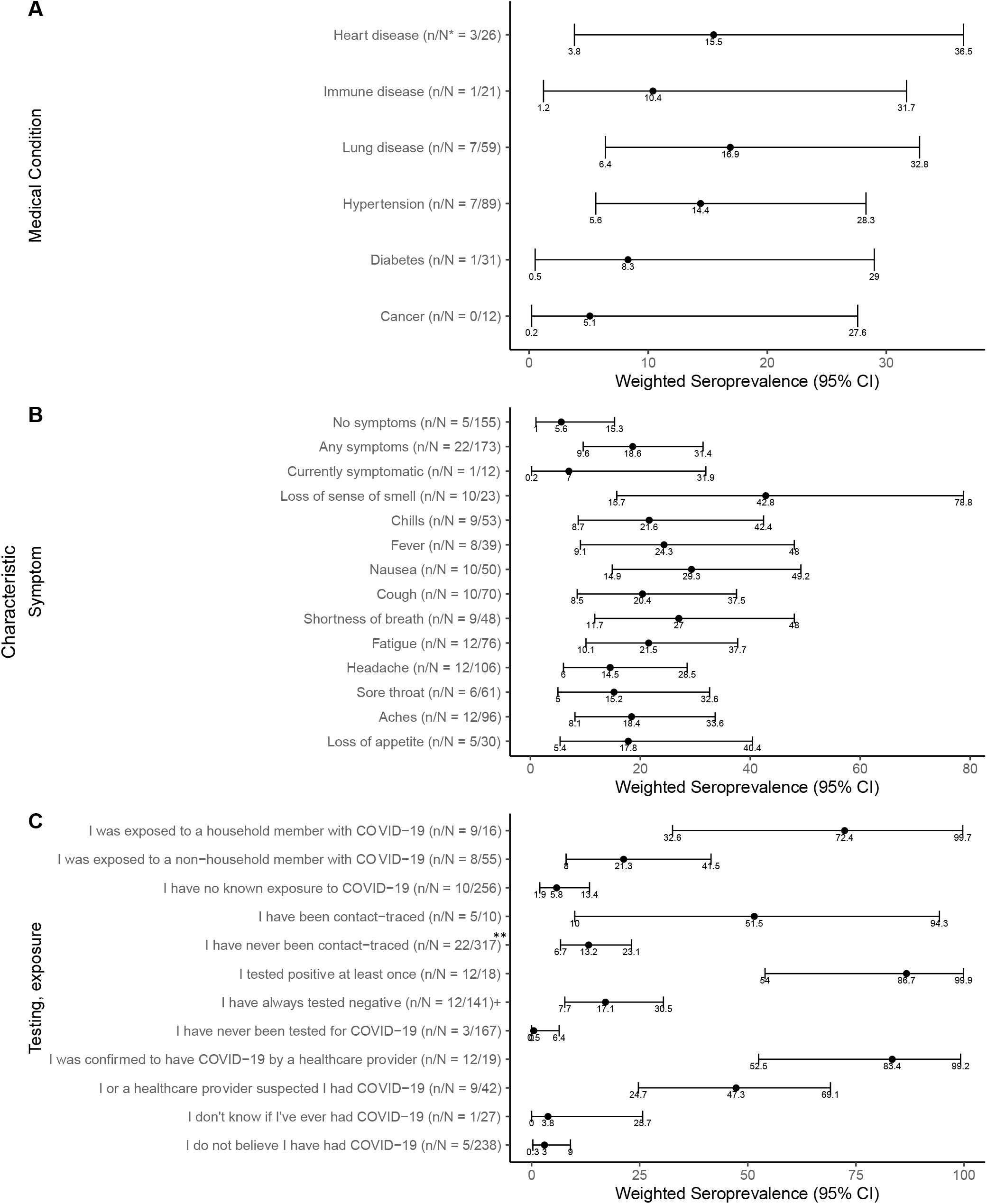
Seroprevalence by Medical, Symptom, Testing and Exposure History. ^*^n refers to the number of individuals that tested positive. N refers to the total number of individuals that responded “Yes” the given question. ^**^being contact-traced refers to individuals that reported being contacted by public health agencies in Massachusetts and told that they were in contact with a person with COVID-19. ^+^Three individuals that did not remember their test results and four individuals that had not received their test results at the time of the survey. Results are for people that reported getting at least one test.

#### Sociodemographic, symptom testing and exposure history by race/ethnicity

Compared to non-Hispanic white individuals, individuals identifying as Hispanic/Latino/Latina were younger and had attained lower education levels (Table 4). They had higher rates of unemployment at the start of the pandemic and were more likely to have their salary impacted by COVID-19. They were more likely to work at a place that offered no benefits such as paid sick leave or work-from-home and were more likely to use the bus as a means of transportation. Housing conditions were also different, with Hispanic/Latino/Latina individuals more likely to live in apartments or condominiums, rent instead of owning their homes, and report a higher density of individuals living in the household.

**Table 4.**
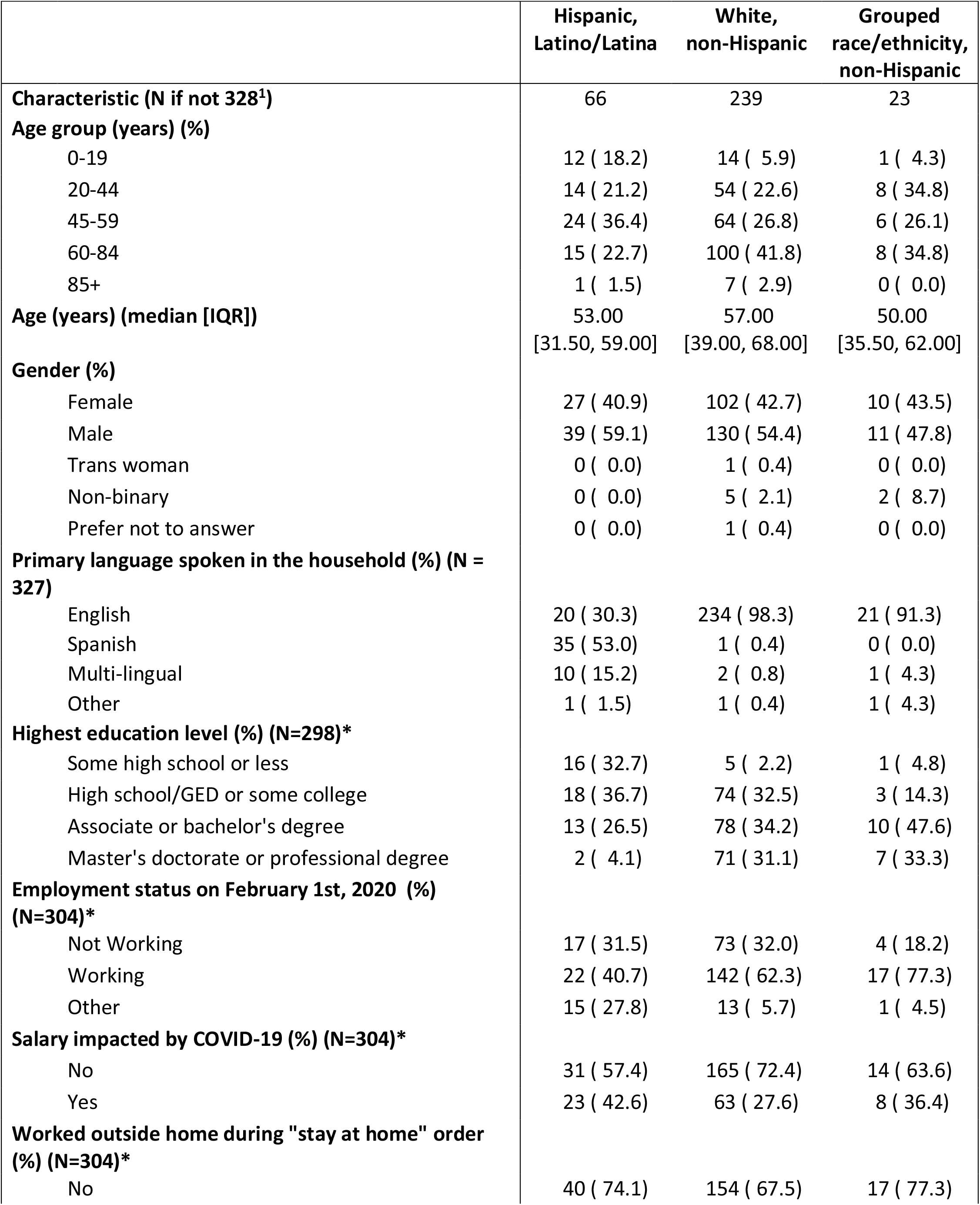
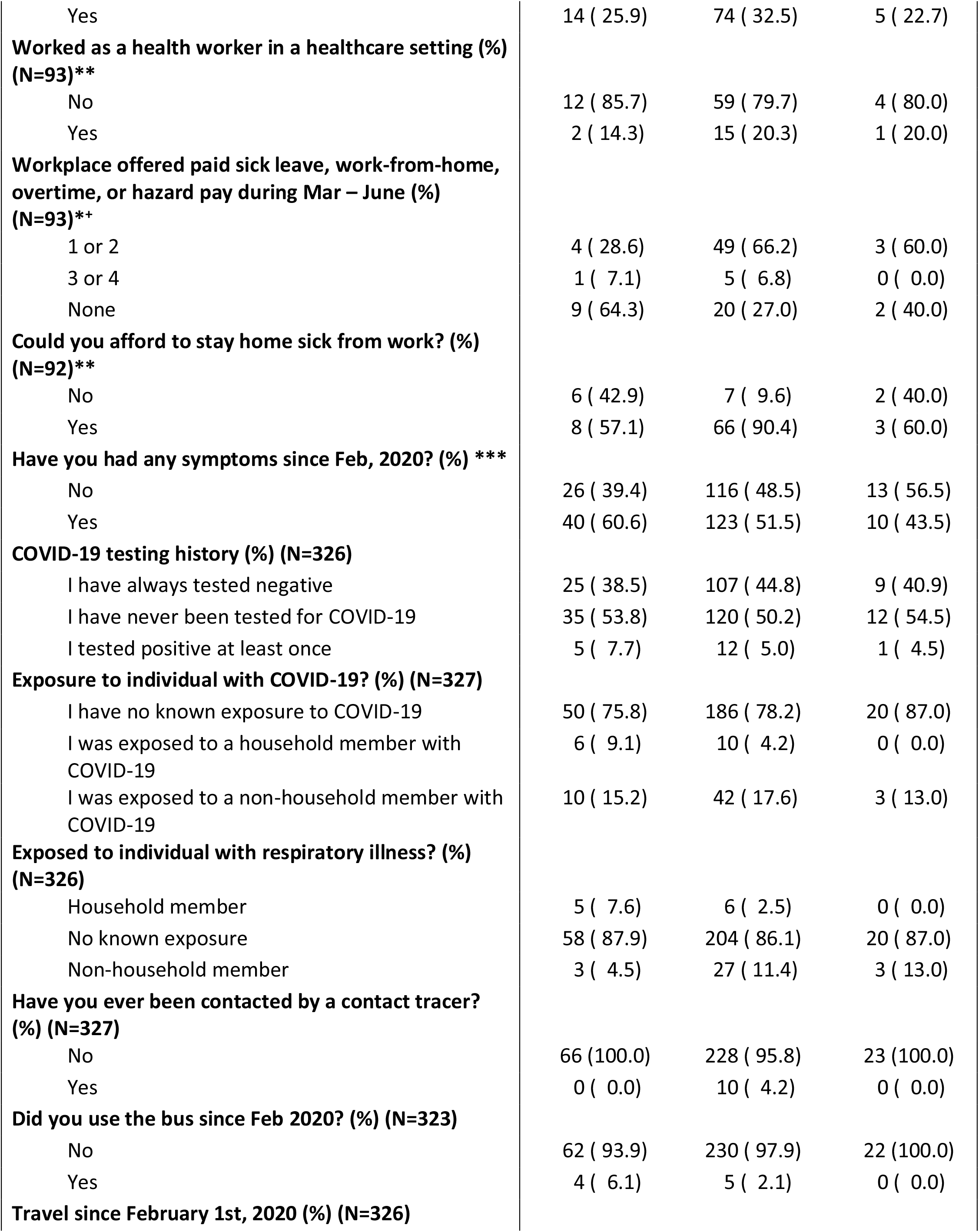
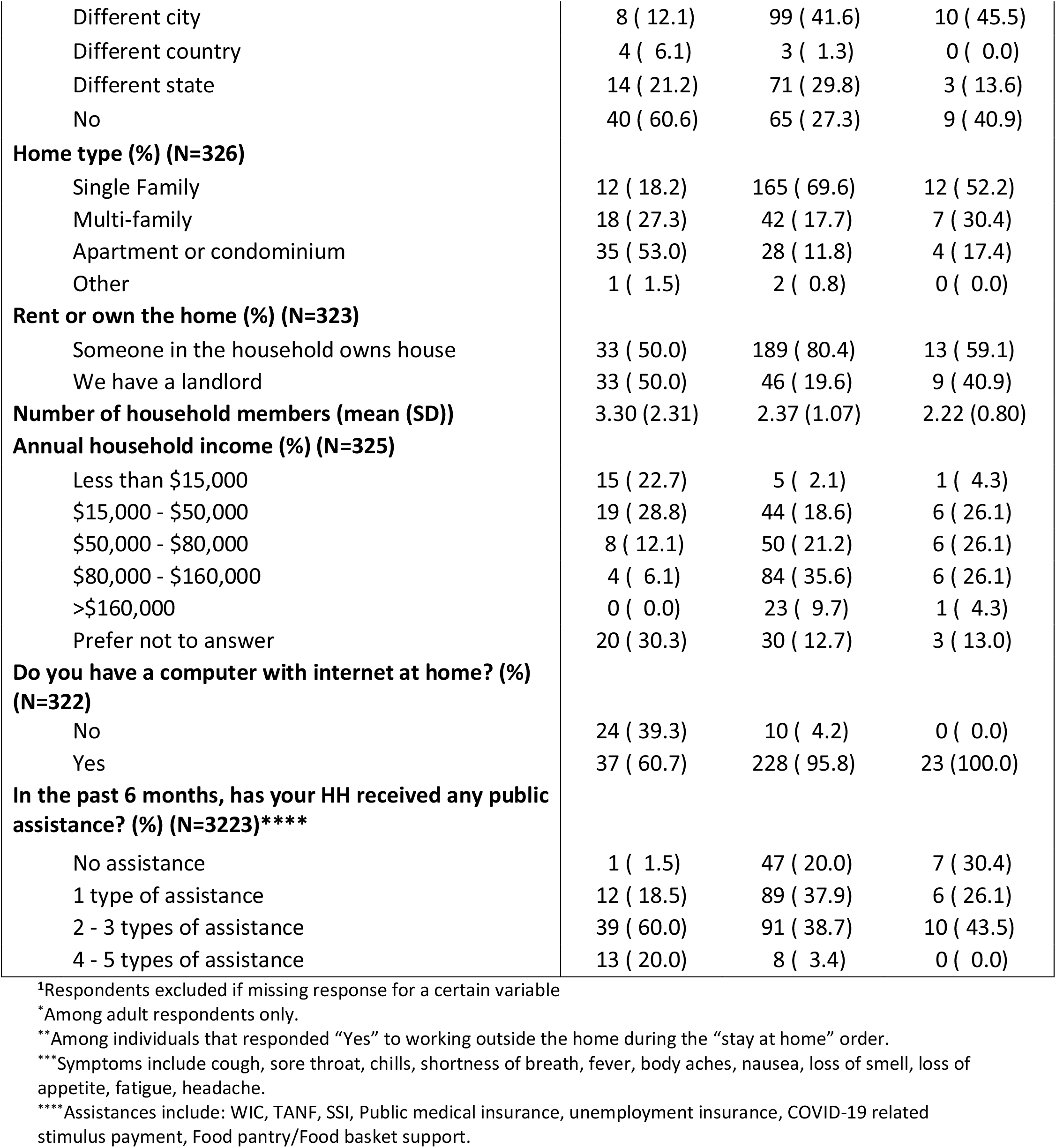
Sociodemographic, symptom, testing and exposure history stratified by race/ethnicity.

### Discussion

We estimated the citywide prevalence of SARS-CoV-2 IgG antibodies to be 13.1% in the City of Holyoke at the end of a second surge of the pandemic in the Commonwealth of Massachusetts between November 2020 and January 2021, prior to widespread vaccination in this community. We identified several groups at high risk of prior infection, on the basis of an elevated risk for SARS-CoV-2 antibodies, which could be targeted for COVID-19-related public health interventions.

In April 2020, a serosurvey using convenience sampling of asymptomatic individuals in the predominantly Hispanic community of Chelsea, MA, demonstrated a seroprevalence of 31.5%.^17^ This serosurvey was limited by non-representative convenience sampling that likely resulted in a biased estimate, and use of a rapid lateral flow immunoassay. Later, between July and August 2020, a university-related population and their household members in Massachusetts demonstrated a lower seroprevalence of 4 - 5.3%.^18^ Our findings in this study conducted several months later are consistent with the increasing number of reported cases during the second surge of COVID-19 in MA. However, the seroprevalence measured in this study was not as high as would be expected approximately ten months into the pandemic. This may be in part due to the impact of decaying antibody titers, the kinetics of which vary depending on the population.^19^

Individuals identifying as Hispanic or Latino/Latina had a higher seroprevalence estimate compared to those identifying as non-Hispanic white, suggesting that these members of the community were at high risk of SARS-CoV-2 infection. Although confidence intervals around effect estimates were wide, this finding is consistent with myriad prior studies documenting racial and ethnic disparities in SARS-CoV-2 metrics affecting minoritized communities nationally and in Massachusetts, including disparities in testing, infections, hospitalizations, and deaths.^20,21^ There are multiple potential mediators of these disparities, lending support to a true difference in risk. In Holyoke, most Hispanic/Latino/Latina communities live in census tracts characterized by high socioeconomic deprivation.^15^ In our study population, compared to non-Hispanic white individuals, individuals identifying as Hispanic/Latino/Latina reported lower education levels, higher unemployment rates, have lower access to benefits such as paid sick leave or work-from-home, and more likely to live in high-density housing. Larger seroprevalence studies are needed to identify the key mediators of the relationship between race/ethnicity and risk of SARS-CoV-2 infection, which can then inform public health interventions tailored to these populations.

Interestingly, individuals from predominantly Spanish-speaking households, almost all who identified as Hispanic/Latino/Latina, had a seroprevalence estimate higher than individuals from English-speaking households. We are not aware of data regarding the availability of language concordant public health messaging and outreach in this community; however, our experience suggests this was limited in Massachusetts, especially during the early phases of the pandemic. The absence of linguistically concordant public health interventions may directly impact an individual’s ability to understand and apply preventive guidance, and thus mediate SARS-CoV-2 infection risk.^22^ Larger and longitudinal serosurveys among populations at high risk are needed to better quantify the magnitude of these relationships.

With regards to the relationship between seropositivity and SARS-CoV-2 exposure history, this study corroborates recent findings. Seroprevalence was higher among individuals with a COVID-19 exposure that was a household member compared to an exposure to a non-household member. This finding highlights the greater importance of intra-household exposures compared to exposure to non-household members.^23^ Given the role of intra-household transmission, public health interventions and resources should be targeted to preventing transmission in the household, such as providing isolation and quarantine sites outside the home, appropriate PPE for individuals taking care of sick family members, timely and sequential testing for exposed household members, and guidance on how to safely distance within the home.

Our findings also identify an important testing gap. Overall, the prevalence of any anti-SARS-CoV-2 antibodies (measured by IgG or IgM) in this survey corresponds to a cumulative case count of 5,593 compared to the city’s actual case count of 3,963 on January 28^th^, 2021 based on testing by RT-PCR.^24^ By this estimate, nearly one third of all SARS-CoV-2 infections in Holyoke were undetected by existing surveillance and screening mechanisms. This level of underascertainment is lower than that demonstrated in other serosurveys throughout the US, a finding that may be explained by the high availability of testing throughout the City of Holyoke, where two public testing sites were established.^25,26^ However, this discrepancy highlights an important testing gap that should be addressed as we prepare for subsequent outbreaks of SARS-CoV-2, future pandemics and other diseases of public health importance.

Our findings also provide potential insight into community-level immunity. Our understanding of immunity following SARS-CoV-2 infection is limited. We do not yet know what level of protection antibodies against SARS-CoV-2 following natural infection provide against subsequent infection or how emerging variants may impact this. It is likely that immunity is more complex than simply the presence of antibodies and relies on other immune-mediating mechanisms such as long-term memory B-cell and T-cell responses. However, the low prevalence of antibodies in this population suggests that at the time of the study, many individuals in the city were vulnerable to infection with SARS-CoV-2. This vulnerability can now be immediately addressed by vaccination. Yet as of May 2021, vaccination rates in Hampden County, MA where Holyoke is located were the lowest in the state.^27^ Furthermore, as of August 31^th^ in Holyoke the percentage of fully vaccinated individuals identifying as Hispanic/Latino/Latina, the group noted to have a high prevalence of SARS-CoV-2 antibodies, was only 32.8%, compared to 59.3% of the non-Hispanic white population.^28^ As such, an emphasis on the rapid delivery of SARS-CoV-2 vaccines to this city is paramount, with a focus on the most-impacted communities. As vaccination progresses, seroepidemiological surveys conducted at the local level will continue to provide key insights by monitoring the prevalence of antibodies as markers of vaccination and natural infection, identifying high-risk groups or areas of low-seroprevalence where vaccination should be focused, and allowing for nuanced and local public health efforts.

This study has several limitations. First, the small sample size led to estimates with wide 95% credible intervals, precluding us from formally comparing seroprevalence between groups and conducting multivariable analyses. Second, our analysis does not account for waning antibody levels, which decay over time.^11^ Third, because we did not interview individuals that declined to participate or did not respond to survey invitation, it is possible that non-response bias may be affecting our findings, though we proactively followed up with households that did not respond to the study via telephone calls and home visits. Fourth, this study was conducted using a single assay. Several studies have shown that variation in the sensitivity and specificity of the serologic test used in a serosurvey can affect seroprevalence estimates.^29^ To limit the effect of this, we used a test that had previously been validated in Massachusetts and conducted a sensitivity analysis using other plausible test sensitivity and specificity values. Finally, we excluded nursing homes and congregate living facilities from this study. Holyoke has a high density of such facilities, and early on during the pandemic suffered a large outbreak at one of these.^30^ However, our goal was to assess community-level epidemiology, as community transmission seems to now be the major driver of the pandemic.

### Conclusion

In conclusion, in this SARS-CoV-2 serosurvey of Holyoke, Massachusetts, between November 2020 – January 2021, antibody prevalence was higher than prior serosurveys in the region, but far from accepted thresholds of “herd immunity.” Risk of infection was high among the Hispanic/Latino/Latina community, the same community with the lowest SARS-CoV-2 vaccination rates in the county. A high prevalence of SARS-CoV-2 antibodies was also noted among other subgroups such as Spanish-speakers and those with household exposures to COVID-19. These differences in infection risk documented through the first year of the pandemic will likely continue absent the necessary locally-focused public health response efforts.

Though national serosurveys provide insights regarding the evolution of the COVID-19 pandemic overall, their scale often precludes an understanding of local details, which are key in pandemic response given the diverse nature of outbreaks throughout the country and the need for local response that matches local issues. City-level serosurveys can provide local authorities with a clearer understanding of disparate viral transmission at a granular level and identify areas of focus to optimize a community-tailored public health response. As we deploy public health efforts to prevent COVID-19, local serosurveys should be implemented more widely. In Holyoke, MA – and in other areas of the US – disparities in SARS-CoV-2 risk must be addressed through proactive public health interventions.

## Data Availability

All data produced in the present study are available upon reasonable request to the authors

## Acknowledgments

We would like to thank other members of the Holyoke Board of Health for their logistical and administrative support with this study. We would also like to thank Alex Morse and Mike Bloomberg for their support and advocacy, Miriam Kwarteng-Siaw for contributing to the implementation of the study and Joseph Rhatigan and the Brigham and Women’s Hospital Division of Global Health Equity for supporting WRM and CPN while in Holyoke. We would like to acknowledge Aaron G. Schmit at the Ragon Institute of MGH, MIT and Harvard for supplying the SARS-CoV-2 RBD protein and monoclonal antibodies.

## References

1. Wu SL, Mertens AN, Crider YS, et al. Substantial underestimation of SARS-CoV-2 infection in the United States. Nat Commun. 2020;11(1):4507. doi:10.1038/s41467-020-18272-4

2. Angulo FJ, Finelli L, Swerdlow DL. Estimation of US SARS-CoV-2 Infections, Symptomatic Infections, Hospitalizations, and Deaths Using Seroprevalence Surveys. JAMA Netw Open. 2021;4(1):e2033706. doi:10.1001/jamanetworkopen.2020.33706

3. Chen X, Chen Z, Azman AS, et al. Serological evidence of human infection with SARS-CoV-2: a systematic review and meta-analysis. Lancet Glob Health. 2021;0(0). doi:10.1016/S2214-109X(21)00026-7

4. Havers FP, Reed C, Lim T, et al. Seroprevalence of Antibodies to SARS-CoV-2 in 10 Sites in the United States, March 23-May 12, 2020. JAMA Intern Med. 2020;180(12):1576. doi:10.1001/jamainternmed.2020.4130

5. Lemieux JE, Siddle KJ, Shaw BM, et al. Phylogenetic analysis of SARS-CoV-2 in Boston highlights the impact of superspreading events. Science. 2021;371(6529). doi:10.1126/science.abe3261

6. Holyoke, M. & Massachusetts | Data USA. Data USA (2018). Holyoke, M. & Massachusetts. Accessed June 4, 2021. https://datausa.io/profile/geo/holyoke-ma/?compare=massachusetts

7. State COVID-19 data show Holyoke, Northampton hardest-hit area communities. Daily Hampshire Gazette. Published April 23, 2020. Accessed June 8, 2021. https://www.gazettenet.com/Cases-of-COVID-19-by-city-and-town-34032621

8. Coronavirus Hot Spots: The 50 Mass. Communities With the Highest Rates of Cases. NBC Boston. Accessed June 8, 2021. https://www.nbcboston.com/news/coronavirus/coronavirus-hot-spots-the-50-mass-communities-with-the-highest-rates-of-cases/2123761/

9. Iyer AS, Azman AS, Bouhenia M, et al. Dried Blood Spots for Measuring Vibrio cholerae-specific Immune Responses. PLoS Negl Trop Dis. 2018;12(1):e0006196. doi:10.1371/journal.pntd.0006196

10. Valentine-Graves M, Hall E, Guest JL, et al. At-home self-collection of saliva, oropharyngeal swabs and dried blood spots for SARS-CoV-2 diagnosis and serology: Post-collection acceptability of specimen collection process and patient confidence in specimens. PloS One. 2020;15(8):e0236775. doi:10.1371/journal.pone.0236775

11. Iyer AS, Jones FK, Nodoushani A, et al. Persistence and decay of human antibody responses to the receptor binding domain of SARS-CoV-2 spike protein in COVID-19 patients. Sci Immunol. 2020;5(52). doi:10.1126/sciimmunol.abe0367

12. Wiens KE, Mawien PN, Rumunu J, et al. Seroprevalence of Severe Acute Respiratory Syndrome Coronavirus 2 IgG in Juba, South Sudan, 2020. Emerg Infect Dis. 2021;27(6). doi:10.3201/eid2706.210568

13. Slater DM. Preparation of Elutions from Dried Blood Spots for ELISAs. protocols.io. doi:10.17504/protocols.io.bsrnnd5e

14. Stringhini S, Wisniak A, Piumatti G, et al. Seroprevalence of anti-SARS-CoV-2 IgG antibodies in Geneva, Switzerland (SEROCoV-POP): a population-based study. The Lancet. 2020;396(10247):313–319. doi:10.1016/S0140-6736(20)31304-0

15. Centers for Disease Control and Prevention/Agency for Toxic Substances and Disease Registry/Geospatial Research, Analysis, and Services Program. CDC/ATSDR Social Vulnerability Index 2018 Database, Massachusetts. https://www.atsdr.cdc.gov/placeandhealth/svi/data_documentation_download.html. Accessed on 6/18/2021.

16. Lumley T. Survey: Analysis of Complex Survey Samples.; 2020. Accessed May 6, 2021. https://CRAN.R-project.org/package=survey

17. Naranbhai V, Chang CC, Beltran WFG, et al. High Seroprevalence of Anti-SARS-CoV-2 Antibodies in Chelsea, Massachusetts. J Infect Dis. 2020;222(12):1955–1959. doi:10.1093/infdis/jiaa579

18. Snyder T, Ravenhurst J, Cramer EY, et al. Serological surveys to estimate cumulative incidence of SARS-CoV-2 infection in adults (Sero-MAss study), Massachusetts, July– August 2020: a mail-based cross-sectional study. BMJ Open. 2021;11(8):e051157. doi:10.1136/bmjopen-2021-051157

19. Post N, Eddy D, Huntley C, et al. Antibody response to SARS-CoV-2 infection in humans: A systematic review. PloS One. 2020;15(12):e0244126. doi:10.1371/journal.pone.0244126

20. Dryden-Peterson S, Velásquez GE, Stopka TJ, Davey S, Lockman S, Ojikutu BO. Disparities in SARS-CoV-2 Testing in Massachusetts During the COVID-19 Pandemic. JAMA Netw Open. 2021;4(2):e2037067. doi:10.1001/jamanetworkopen.2020.37067

21. Mackey K, Ayers CK, Kondo KK, et al. Racial and Ethnic Disparities in COVID-19–Related Infections, Hospitalizations, and Deaths. Ann Intern Med. 2020;174(3):362–373. doi:10.7326/M20-6306

22. Maleki P, Al Mudaris M, Oo KK, Dawson-Hahn E. Training Contact Tracers for Populations With Limited English Proficiency During the COVID-19 Pandemic. Am J Public Health. 2021;111(1):20–24. doi:10.2105/AJPH.2020.306029

23. Yu H-J, Hu Y-F, Liu X-X, et al. Household infection: The predominant risk factor for close contacts of patients with COVID-19. Travel Med Infect Dis. 2020;36:101809. doi:10.1016/j.tmaid.2020.101809

24. Archive of COVID-19 Weekly Public Health Reports | Mass.gov. Accessed May 7, 2021. https://www.mass.gov/info-details/archive-of-covid-19-weekly-public-health-reports

25. Menachemi N, Yiannoutsos CT, Dixon BE, et al. Population Point Prevalence of SARS-CoV-2 Infection Based on a Statewide Random Sample - Indiana, April 25-29, 2020. MMWR Morb Mortal Wkly Rep. 2020;69(29):960–964. doi:10.15585/mmwr.mm6929e1

26. Anand S, Montez-Rath M, Han J, et al. Prevalence of SARS-CoV-2 antibodies in a large nationwide sample of patients on dialysis in the USA: a cross-sectional study. Lancet Lond Engl. 2020;396(10259):1335–1344. doi:10.1016/S0140-6736(20)32009-2

27. Weisman R. In state’s least immunized county, vaccine holdouts remain as wariness persists-The Boston Globe. Boston Globe. https://www.bostonglobe.com/2021/05/22/metro/states-least-immunized-county-vaccine-holdouts-remain-wariness-persists/. Published May 22, 2021. Accessed June 18, 2021.

28. COVID-19 Vaccine Tracker. Pioneer Institute. Published April 16, 2021. Accessed September 11, 2021. https://pioneerinstitute.org/covid/covid-19-vaccine-tracker/

29. Fotis C, Meimetis N, Tsolakos N, et al. Accurate SARS-CoV-2 seroprevalence surveys require robust multi-antigen assays. Sci Rep. 2021;11(1):6614. doi:10.1038/s41598-021-86035-2

30. The COVID-19 Outbreak at the Soldiers’ Home in Holyoke. An Independent Investigation Conducted for the Governor of Massachusetts.; 2020:174.

