## Supplementary Material for "Disparities in SARS-CoV-2 exposure: evidence from a citywide seroprevalence study in Holyoke, Massachusetts, USA"

### Supplement: Holyoke Seroprevalence Study

#### 1 Bayesian model

##### 1.1 Setup

Following the approach in Stringhini et al. (2020), we use a Bayesian logistic regression model with a random intercept for household clustering. We account for the sensitivity and specificity of the antibody test.

$$
\begin{aligned}
x_i &\sim Bernoulli(p_i * \theta^+ + (1-p_i) * (1-\theta^-)) \\
logit(p_i) &= \alpha_h + \beta_0 + \mathbf{Z}_i\boldsymbol{\beta} \\
\alpha_h &\sim Normal(0, \sigma^2) \\
x^+ &\sim Binomial(n^+, \theta^+) \\
x^- &\sim Binomial(n^-, 1-\theta^-)
\end{aligned}
$$

where $x_i$ is the result of the particular antibody test and $p_i$ is the true underlying probability for the $i$th person. The sensitivity, $\theta^+$ is determined by $n^+$ RT-PCR positive controls from the lab validation study with $x^+$ testing positive (true positive). Similarly, the specificity, $\theta^-$ is determined by $n^-$ pre-pandemic controls with $x^-$ testing negative (true negative). The random intercept $\alpha_h$ is indexed by household $h$. Finally, $\mathbf{Z_i}$ is the collection of covariates we will use for weighting, including age, sex, race ethnicity, and social vulnerability index. This model was implemented using the `rstan` package in R with 5,000 iterations and four chains with 250 warm-up and convergence was assessed using the R-hat statistic.

##### 1.2 Prior distributions

The prior distributions for $\theta^+$ and $\theta^-$ were flat from 0 to 1, equivalent to Uniform(0,1). We used lighter tailed priors for the $\beta$ coefficients and $\sigma$ because the No-U-Turn Sampler implemented in `rstan` may have convergence issues with heavy-tailed priors (Ghosh et al., 2018). Notably, we did experience convergence issues when trying Cauchy priors for the $\beta$ parameters and a flat prior for $\sigma$. For the $\beta_0$ intercept, we utilized a Normal(0,10) prior. For other $\beta$ parameters, a Normal (0,2.5) or Normal(0,5) was employed – the latter was used for covariates that were expected to have profound differences in seroprevalence between levels (e.g. occurrence of COVID-19 symptoms, known COVID-19 exposure, suspected or confirmed COVID-19 disease) to ensure we did not push the odds ratios between groups towards null. For $\sigma$, we used a truncated Normal prior with mean 1 and standard deviation 1; notably, we tried a half-Normal prior (mean 0) with a standard deviation of 1, with no impact on results.

##### 1.3 Weighting

We first construct population-representative weights based on $V$ categorical variables. In our case, we have $V = 4$ variables: age, race ethnicity, sex, and social vulnerability index (SVI). Subgroups will be denoted based on their variable and level indexed by $v$ and $l$, respectively. For example, let $V = 1$ correspond to the age variable and $L = 1$ denote persons aged 0-19 years.

Consider partitioning individuals based on each variable into distinct subgroups indexed by $j$. Let $\mathcal{J}$ be a collection of all distinct subgroups and $\mathcal{I}_j$ denote the collection of indices for all respondents within the distinct subgroup $j$ (e.g. Hispanic females aged 0-19 years living in a high SVI area).

Let $\mathcal{J}_{vl}$ indicate the collection of all distinct subgroups $j$ containing individuals with one common characteristic indicated by variable $v$ and level $l$. Let $\mathcal{I}_{vl}$ be the collection of indices for all respondents who have the chosen characteristic indicated by variable $v$ and the level $l$ (e.g. $\mathcal{I}_{11}$ would contain the indices of all persons aged 0-19 years in our sample).

The weights were calculated in the `survey` package in R using raking. Raking is an iterative procedure that assigns weights to each individual, $w_i$, based on the marginal proportions for each variable and level such that:

$$\sum_{i}^{n} w_i = N$$

$$\sum_{i \in I_{vl}} w_i = N_{vl} \text{ for each variable } v \text{ and level } l$$

where $n$ is the sample size, $N$ is the size of the target population, and $N_{vl}$ is the population size of the subgroup $v, l$ in the target population. For example, $N_{11}$ would be the number of persons aged 0-19 years in Holyoke. Note that raking does not necessarily ensure that $\sum_{i \in I_j} w_i = N_j$; this would only be possible if individual-level data on the City of Holyoke was publicly available. Hence, we rely on the marginal distributions from the American Community Survey in 2019 for each variable (**Table 1**).

**Table 1. Holyoke Demographics from American Community Survey 2019**

| Variable | Proportion |
|---|---|
| **Age (years)** | |
| 0-19 | 0.258 |
| 20-44 | 0.357 |
| 45-59 | 0.189 |
| $\geq 60$ | 0.196 |
| **Race ethnicity** | |
| non-Hispanic white | 0.413 |
| Hispanic | 0.539 |
| non-Hispanic other | 0.048 |
| **Sex** | |
| Female | 0.512 |
| Male and other | 0.488 |
| **Social vulnerability index** | |
| Low | 0.222 |
| High | 0.778 |

#### 1.4 Overall prevalence

For each iteration $r$, we take each $\beta_0^r$, $\boldsymbol{\beta}^r$, and $\sigma^r$ and complete the following steps:

**Step 1:** Compute $p_j^r = \int_\alpha expit(\alpha_h^r + \beta_0^r + \mathbf{Z}_j \boldsymbol{\beta}^r) f_\alpha(\alpha_h^r)$ for each subgroup $j \in \mathcal{J}$

**Step 2:** Calculate $p^r = \frac{\sum_{j \in \mathcal{J}} (\sum_{i \in \mathcal{I}_j} w_i) p_j^r}{N}$

Let $\mathbf{Z}_j$ be a binary vector aligning with the relevant subgroup $j$. For Step 1, the integral is computed with a change of variables (see Stringhini paper) and using the `integrate()` function in R. Step 2 approximates the approach used in Stringhini et al. (2020) paper and can be used when cross-tabulations for the target populations are not available. The posterior median (median across all iterations $r$) is the overall prevalence estimate. The 2.5th and 97.5th percentiles is the 95% credible interval for the overall prevalence.

#### 1.5 Subgroup prevalence

For any subgroup corresponding to variable $v$ and level $l$,

**Step 1:** Same as the above

**Step 2:** Calculate $p_j^r = \frac{\sum_{j \in \mathcal{J}_{vl}} (\sum_{i \in \mathcal{I}_j} w_i) p_j^r}{N_{vl}}$

For a subgroup not included as a weighting variable, let $X$ denote a binary variable. For ease of exposition, let's say interest lies in estimating the prevalence among the subgroup where $X = 1$.

**Step 0**: Re-run the Bayesian procedure using the following model,

$$logit(p_i) = \alpha_h + \beta_0 + \mathbf{Z}_i\boldsymbol{\beta} + \gamma X$$

Now, for each iteration $r$, we take each $\beta_0^r$, $\boldsymbol{\beta}^r$, $\gamma^r$, and $\sigma^r$ and complete the following steps:

**Step 1**: Compute probabilities for each distinct weighting subgroup $j$ among persons with $X = 1$,

$$p^r_{j,X=1} = \int_\alpha expit(\alpha_h^r + \beta_0^r + \mathbf{Z}_j\boldsymbol{\beta}^r + \gamma^r) f_\alpha(\alpha_h^r)$$

**Step 2**: Calculate prevalence within subgroup $X = 1$,

$$p^r_{X=1} = \frac{\sum_{j\in\mathcal{J}}(\sum_{i\in\mathcal{I}_j\cap\mathcal{I}_{X=1}} w_i)p^r_{j,X=1}}{\sum_{j\in\mathcal{J}}\sum_{i\in\mathcal{I}_j\cap\mathcal{I}_{X=1}} w_i}$$

where $\mathcal{I}_{X=1}$ are the indices for persons with $X = 1$.

#### 2 Test sensitivity and specificity considerations

Estimates of test specificity were calculated using 1548 pre-pandemic samples from the Boston area. Estimates of test sensitivity were calculated among individuals not hospitalized with COVID-19 disease (n=94). Disease positive individuals had confirmed RT-PCR tests and up to three blood samples sent for SARS-CoV-2 testing at varying time points after symptom onset. We utilized the second time point for all individuals, which was collected at a median of 28 days after symptom onset and ranged between 6 and 90 days.

The table below gives the sensitivity and specificity estimates for each antibody type and combination. Our main measure used the sensitivity and specificity associated with the presence of IgG antibodies.

**Table 2. Sensitivity and specificity estimates used in analyses**

| **Characteristic** | $n^-$ | $x^+_{n^-}$ | **Specificity** | $n^+$ | $x^+_{n^+}$ | **Sensitivity** |
|---:|---|---|---|---|---|---|
| **IgG** | 1548 | 7 | 99.5 (99.0-99.8) | 94 | 67 | 70.6 (61.2-79.3) |
| **IgM** | 1548 | 9 | 99.4 (98.9-99.7) | 94 | 38 | 40.0 (30.0-50.0) |
| **IgG or IgM** | 1548 | 16 | 98.9 (98.3-99.3) | 94 | 71 | 75.0 (65.8-83.2) |
| **IgG and IgM** | 1548 | 0 | 99.9 (99.8-100) | 94 | 34 | 35.7 (26.4-45.7) |

We also performed a sensitivity analysis for the overall prevalence using sensitivity estimates corresponding to five other testing time points: first testing date, first testing date if it occurred at or after 14 days, second testing date (above), second testing date if it occurred at or after 14 days, and the third testing date. Interestingly, sensitivity increases the later the test is performed. The table gives the sensitivity estimates for each of these testing time points:

**Table 3. Alternate sensitivity estimates for IgG antibodies**

| Time point | Median (days) | $n^+$ | $x^+_{n^+}$ | Sensitivity |
|---|---|---|---|---|
| **1** | 20 | 96 | 58 | 60.2 (50.7-69.6) |
| **1 ($\geq$ 14 days)** | 26 | 71 | 53 | 73.7 (63.4-83.3) |
| **2** | 28 | 94 | 67 | 70.6 (61.2-79.3) |
| **2 ($\geq$ 14 days)** | 28.5 | 92 | 67 | 72.1 (62.8-80.8) |
| **3** | 47 | 84 | 67 | 78.8 (69.7-86.6) |

We implemented the Bayesian procedure to estimate overall prevalence for the varying sensitivity estimates presented in Table 3. The results are plotted below.

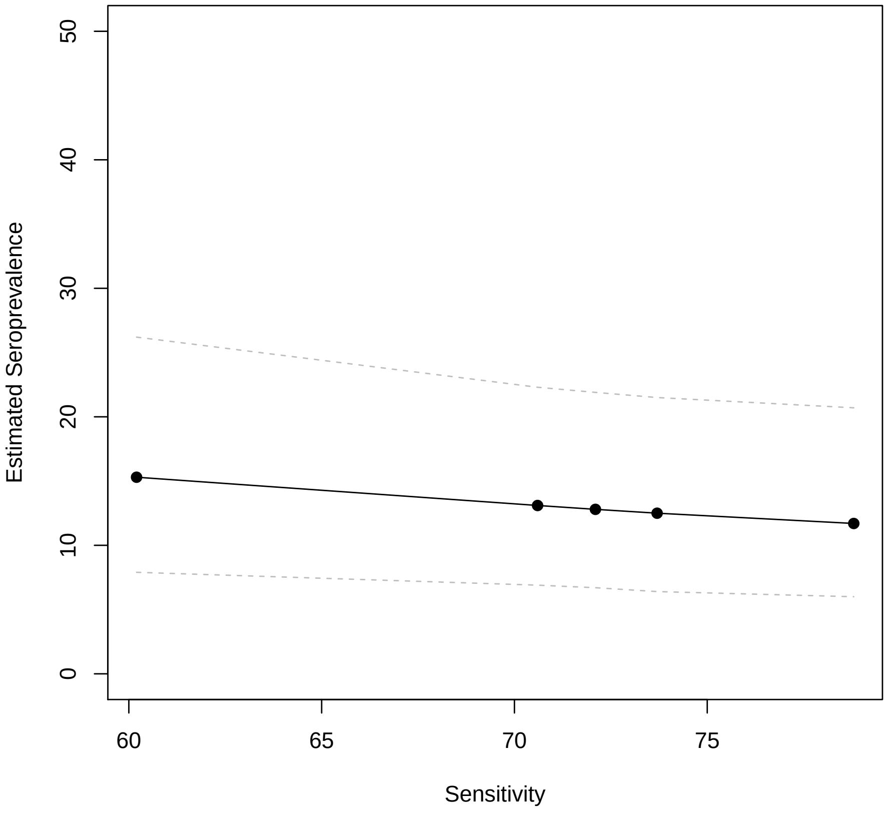
